# Longitudinal analysis of virus load, serum antibody levels and virus neutralizing activity *in vitro* in cases with less severe COVID-19

**DOI:** 10.1101/2020.08.20.20174912

**Authors:** Bertram Flehmig, Michael Schindler, Natalia Ruetalo, Ramona Businger, Manfred Bayer, Angelika Haage, Thomas Kirchner, Karin Klingel, Andrea Normann, Lutz Pridzun, Despina Tougianidou, Michael B. Ranke

**Author notes:** shared first authors (Bertram Flehmig, Michael Schindler). Corresponding author, Prof. Dr. Bertram Flehmig Mediagnost GmbH Aspenhaustrasse 25 72770 Reutlingen (Germany).

## Abstract

**Summary:** *Background:* Patients infected with SARS-CoV-2 exhibit a highly variable clinical course, varying from barely discernible signs of disease, to moderate flu-like symptoms and, occasionally, with life-threatening pneumonia and/or cytokine storm. The relationship between the nasopharyngeal virus load, IgA and IgG antibodies to both the S1-RBD-protein and the N-protein as well the neutralizing activity (NAbs) against SARS-CoV-2 in the blood of moderately afflicted COVID-19 patients has not been investigated longitudinally so far. Methods
Several new serological methods to examine these parameters were developed and validated for the longitudinal investigation in three patients of a family which underwent a mild course of COVID-19.

**Findings:** We observed that the virus load had almost completely disappeared after about four weeks, whereas serum antibodies showed a contrasting course. IgA levels to S1-RBD-protein and, to a lesser extent, to the N-protein, peaked about three weeks after clinical disease onset but declined soon thereafter. IgG levels rose continuously, reaching a plateau approximately six weeks after disease onset. NAbs in serum reached a peak about four weeks after disease onset but dropped to a lower level about six weeks later.

**Interpretation:** Our data establishes associations of virus neutralization and a serological immune response foremost against Sars-CoV-2 S1-RDB-protein in a longitudinal manner.

## Introduction

According to our current understanding, coronavirus SARS-CoV-2, which causes COVID-19,^1,2^ is transmitted among humans via droplets from the respiratory tract. The high level of contagiousness of SARS-CoV-2 is probably related to a high virus load in human sputum, on the one hand, and to the protective effect of proteins against virus material in the respiratory tract on the other. In a recent cell culture study, SARS-CoV-2 was isolated from the saliva of individuals over a period of two weeks after the onset of a symptomatic infection.^3^ The study provides information about the excretion of the virus in the sputum and stool of severely-ill, hospitalized patients. Data from moderately-ill patients, however, is yet unavailable in terms of the length of time over which SARS-CoV-2 is excreted into the respiratory tract of these patients. In addition, there is limited information about the duration of virus production in the upper respiratory tract and the amount of virus in the sputum of such patients. Furthermore, little is known about the course of the immune response with regard to the various types of antibodies against SARS-CoV-2 proteins and their virus neutralizing potential.

Here we report on the extent of SARS-CoV-2 RNA present in nasal and oro-pharyngeal fluid as well as on the level of IgG and IgA antibodies against the S1-RBD-protein and the N-protein of SARS-CoV-2 in blood and their ability to neutralize SARS-CoV-2 *in vitro*. The samples from three patients belonging to the same family were examined over a period of 15 weeks, starting with the onset of the disease.

## Patients and Methods

### Ethics

Experiments conformed to the Declaration of Helsinki Principles. Written informed consents were obtained from all blood donors prior to the study. The study was approved by the Independent Research Ethics Committee (IEC) of the University of Tuebingen, IEC-Project Number 672/2020A

### Patients

All patients are related and live in the same household. *Patient 1:* Adult male. Coughing is the first symptom, lasting until day 8 after onset of disease. Fever (38.1°C) on days 2 and 3. Symptoms on days 2-8 include headache, muscle pain in arms and legs and tiredness. Apart from occasional coughing after day 8, the patient’s well-being improves until full recovery of health is reached. *Patient 2:* Adult female. Coughing is the first symptom, occurring three days after disease onset in patient 1, followed by elevated body temperature (37-38°C) on days 4 and 5. Headache and limb pain occur on days 2-8. Persistent tiredness since day 4 and anosmia from day 5-13, with subsequent full recovery. *Patient 3:* Adult female. Three days after patient 1 she began coughing, had elevated temperature (37-38°C) on days 2 and 3, muscle pain on day 3, rhinitis on days 5-7, and hyposmia on days 2-8. She recovered fully by day 8.

In each of the patients, SARS-CoV-2 RNA had been diagnosed via nasal and oro-pharyngeal fluids, with testing done in a commercial diagnostic laboratory on day 1, when the first symptoms had presented. A quantification of the virus load in these specific samples is not available.

### Methods

In our study, blood samples were taken from patients over more than 100 days at the following time points (t1 – t9): <u>patient 1:</u> on days 13 (t1), 20 (t2), 25 (t3), 33 (t4), 40 (t5) 48 (t6), 60 (t7), 74 (t8) and 109 (t9) after onset of symptoms; <u>patient 2:</u> on days 10 (t1), 17 (t2), 22 (t3), 30 (t4), 37 (t5), 45 (t6), 57 (t7), 71 (t8) and 107 (t9) after experiencing first symptoms; and <u>patient 3:</u> on days 11 (t1), 18 (t2), 23 (t3), 31 (t4), 38 (t5), 46 (t6), 58 (t7), 72 (t8) and 106 (t9) days after onset.

Nasopharyngeal samples were taken from patients at t1-t5, and t9.

### SARS-CoV-2 RT-qPCR^4^

The virus RNA was isolated from swab samples using the High Pure Viral Nucleic Acid Kit (Roche, 11858874001) as follows: each swab was incubated for 10 min at 70°C in 350 μL binding buffer, 50 μL proteinase K and 4 μL poly(A)-RNA. After the addition of 250 μL of phosphate-buffered saline, samples were mixed by vigorous vortexing, after which a 600-μL sample was processed according to the manufacturer’s specifications. Nucleic acids were eluted in 50 μL of elution buffer, 5 μL of the eluted sample was examined in a total RT-qPCR reaction volume of 20 μL, according to the manufacturer’s specifications, using LightCycler 480 (Roche) instrumentation. The detection of SARS-CoV-2 RNA was done by means of the SARS-CoV-2-realDETECT kit (Mediagnost GmbH, Germany). Amplification of the E gene and the human RNase-P control were carried out in multiplex RT-qPCR reactions. A recombinant *in* vitro-transcribed RNA standard (4BLqSARS-CoV 2-RNA, C0301001, 4base lab AG, Germany) encoding the SARS-CoV-2 primer and probe-binding sites described by Corman et al.^4^ and the Centers for Disease Control and Prevention, USA^5^ was applied for quantification.

### Detection of IgA and IgG Antibodies against SARS-CoV-2-S1-RBD-Protein

**IgG Antibody Assay:** The ELISA test system E 111-IVD developed by Mediagnost GmbH, Reutlingen (Germany) was employed according to the manufacturer’s instructions. This test system is a two-step enzyme-linked immunosorbent assay for the detection of IgG antibodies directed against SARS-CoV-2 S1, specifically targeting the receptor binding domain (RBD) protein. The solid phase consists of a 96-well microtiter plate (Greiner, Bio One, Frickenhausen, Germany) that is coated with the recombinant SARS-CoV-2-S1-RBD-protein.

Briefly, the test methodology can be described as follows: antibodies from patients’ serum that are directed against the SARS-CoV-2-S1-RBD-protein bind to the immobilized antigen. Next, a horseradish peroxidase (HRP) conjugated goat anti-human IgG binds to the human IgG antibodies. The following step involves the substrate for the HRP being added, by which the substrate is converted by HRP from colorless into blue; and, after the addition of a stop solution, the color changes to yellow. The extinctions of the yellow solution can be measured at a wavelength of 450 nm, with reference at 620 nm. The lower threshold is determined by using human serum without antibodies against SARS-CoV-2; the cut-off towards positive values was estimated by using the extinction value as the negative control times five. Thus, samples showing extinctions that were five times higher than the negative control can be interpreted as being positive for anti-SARS-CoV-2 S1. Increasing extinctions represent increasing amounts of antibodies to SARS-CoV-2 S1 protein. The specificity was substantiated by means of sera containing high levels of antibodies against other viral diseases - such as the hepatitis viruses A, B and C, Epstein Barr virus, cytomegalovirus, influenza viruses A and B, varicella zoster virus and human herpes simplex virus type 1 - as determined at the Institute for Medical Virology and Epidemiology and the Institute for Pathology and Neuropathology of the University of Tuebingen, Germany. The results for each of the tested sera were negative in terms of antibodies against the SARS-CoV-2 S1 protein. Serum from patients infected with other human coronaviruses such as HKU-1 and NL63 also proved to be negative.

**IgA Antibody Assay:** The ELISA test system for the detection of IgA antibodies to SARS-CoV-2 S1-RBD protein was analogous to the anti-SARS-CoV-2 IgG ELISA, except that the HRP-labeled detection antibody was directed against human IgA antibodies.

IgA and IgG assays for the determination of antibodies to the SARS-CoV-2 nucleoprotein (N-protein) in sera were developed in analogy to the assays for anti-SARS-CoV-2-S1-RBD-protein as described above. The first step in these assays involve the SARS-CoV-2 N-protein being bound to the solid phase.

### Virus Neutralization Assay (VNA)

By means of the VNA, we quantified *in vitro* the ability of human sera from patients to inhibit the infection of human cells Caco-2 (human colorectal adenocarcinoma) with a strain of SARS-CoV-2 virus. The SARS-CoV-2 strain icSARS-CoV-2-mNG^6^ was obtained from the World Reference Center for Emerging Viruses and Arboviruses (WRCEVA) of the UTMB (University of Texas Medical Branch). To generate icSARS-CoV-2-mNG stocks, Caco2 cells were infected, the supernatant was harvested 48 hours post infection, centrifuged and stored at -80°C. For MOI (multiplicity of infection) determination, a titration using serial dilutions of the virus stock was conducted. The number of infectious virus particles per ml was calculated as the (MOI ^x^ cell number)/(infection volume), where MOI = -ln (1 - infection rate).

For neutralization experiments, 1 x10^4^ Caco-2 cells/well were seeded in 96-well plates the day before infection, in media containing 5% FCS. Cells were infected with the SARS-CoV-2 strain icSARS-CoV-2-mNG at a MOI=1or mock-infected. Immediately after infection, serum for COVID-19 patients was added to each well in two-fold serial dilutions from 1:40 up to 1:5120. The cells were fixed 48h post infection with 2% PFA and stained with Hoechst 33342 (1 μ g/mL final concentration) for 10 minutes at 37°C. The staining and fixing solution was then removed, after which PBS was added. In order to quantify the infection rates, images were taken with the Cytation3 (Biotek Hoechst) and mNG+ cells were automatically counted by the Gen5 Software (Biotek). Titers of neutralizing antibodies (NAbs) were calculated as the half-maximal inhibitory dose (ID50) using 4-parameter nonlinear regression (GraphPad Prism).

## Results

The results shown here represent the mean of two independent test series. The numbers of SARS-CoV-2 RNA copies in swab samples observed over approximately five weeks, starting with the onset of COVID-19 symptoms in our patients, are illustrated in Figure 1.

**Figure 1:**
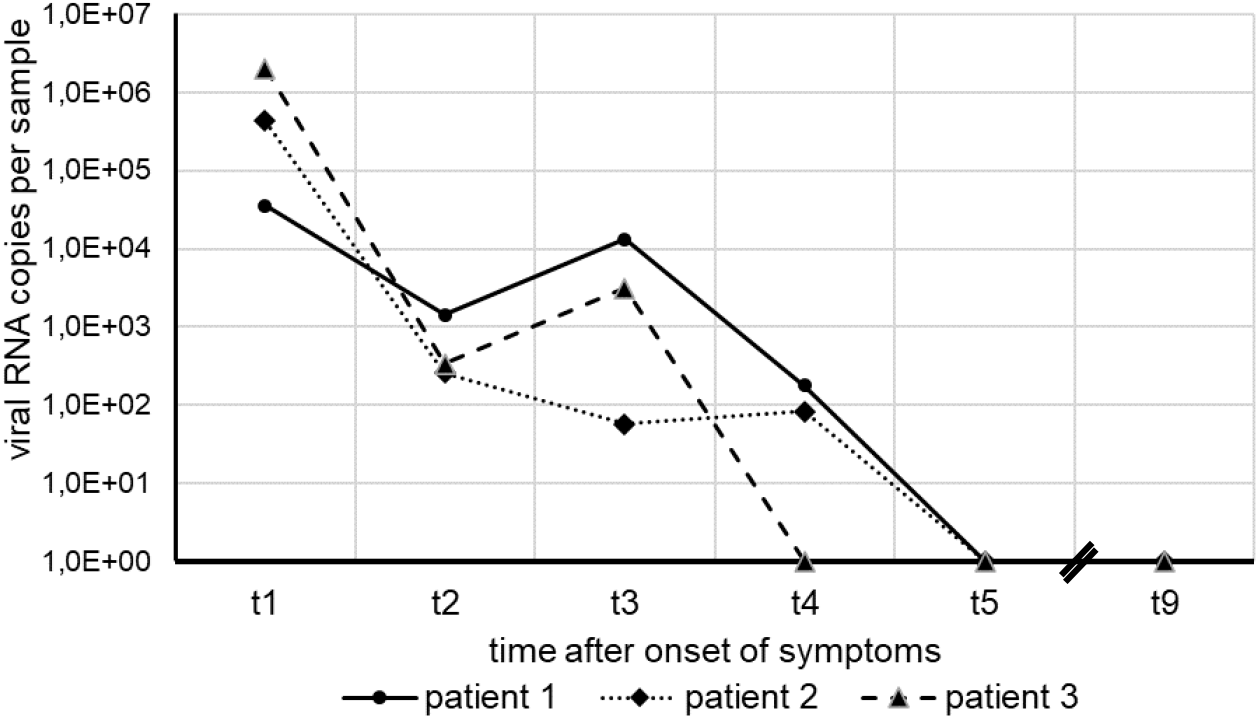
Time course of SARS-CoV-2 RNA detection in nasal and oro-pharyngeal swabs of three clinically ill patient.

In each of the patients, the amount of virus RNA was highest about one week after the onset of symptoms, with a decline of several orders of magnitude following thereafter. Within a short period of 31 days (patient 3), 38 days (patient 2) and 40 days (Patient 1), virus RNA was no longer detectable in nasal and oro-pharyngeal fluid. All sera taken from the patients revealed negative RT-qPCR results for SARS-CoV-2 RNA, suggesting the absence of viremia.

The results of the levels of anti-SARS-CoV-2 S1-RBD antibodies and the anti-SARS-CoV-2 N antibodies - IgA and IgG - expressed in terms of the absorbance (see Methods), and the titers of the virus neutralizing antibodies (NAbs) expressed as the half-maximal inhibitory dose (ID50) during the observation phase, are illustrated in Figures 2a-f.

**Figure 2:**
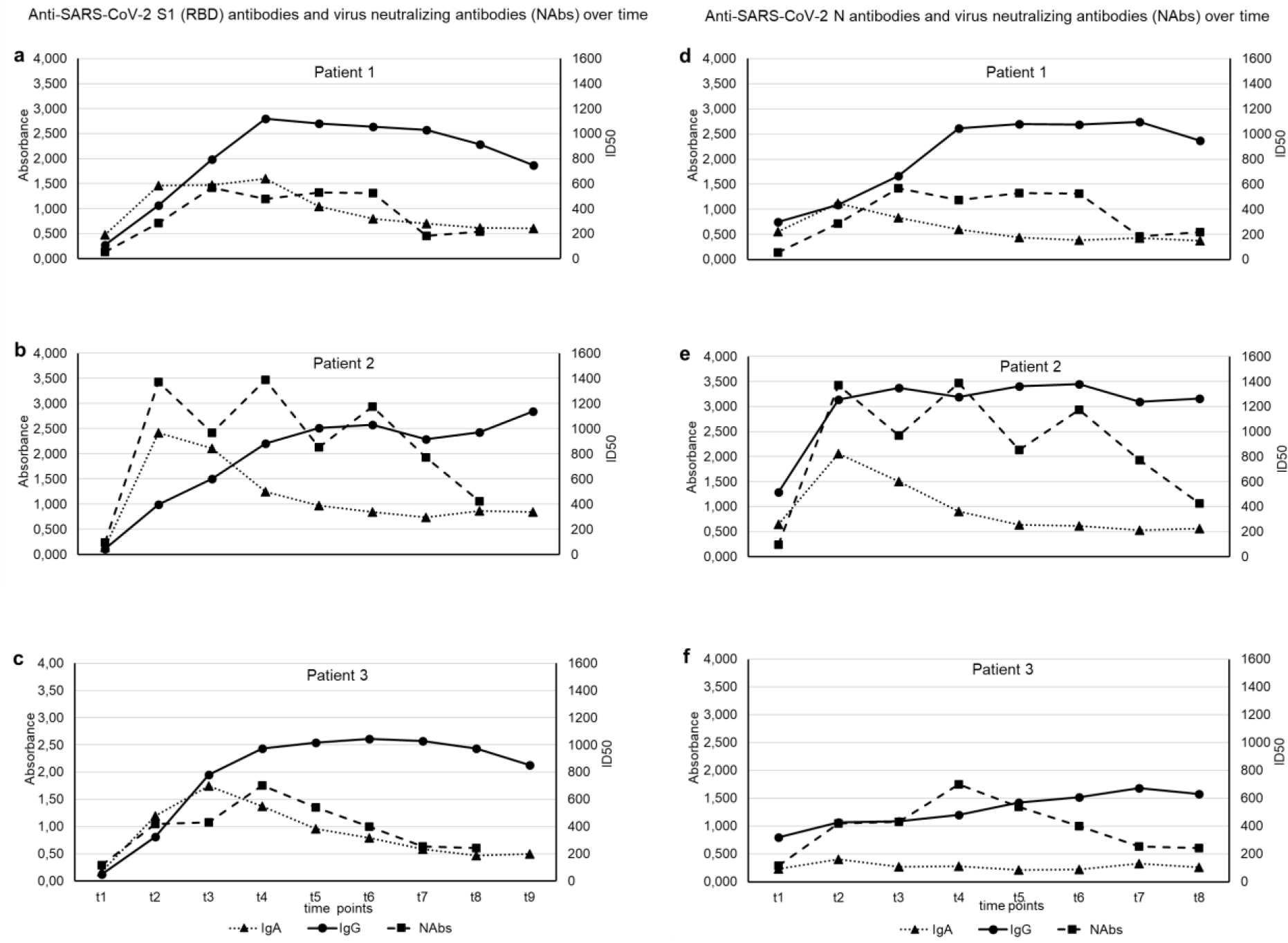
Time course of anti-SARS-CoV-2 S1-RBD (Fig.2 a-c) and anti-SARS-CoV-2-N (Fig. 2 d-f) IgG, IgA and neutralizing antibody development of three clinically ill patients (patient 1, patient 2 and patient 3) using sera taken at different days (t1 – t9) after the onset of COVID-19 disease.

The course of the antibodies (Abs) against the two different regions of SARS CoV-2, the S1-RBD protein and the N-protein and the NAbs in the three affected individuals show a characteristic pattern. All patients exhibited an early rise in the IgA Abs against S1-RBD-protein and N-protein, which peaked around three weeks (t2-t3) and subsequently declined to basal levels at around five weeks (t5). Quantitatively, the response of IgA-Abs was most profound in patient 2, who also had the most severe clinical course. Changes in IgG Ab levels against S1-RBD-protein or N-protein were qualitatively similar but showed a different dynamic. While anti-S1-RBD IgG-Abs rose slowly from the beginning, reaching a maximum at around five weeks after disease onset (t4-t5), they remained elevated at a high level,whereas the anti-N IgG-Abs had already increased at around two weeks (t1) in all patients and rose further thereafter. Again, patient 2 showed the highest levels. The course of N-Abs in the patients was time-dependent and qualitatively similar in both types of Abs against the virus S1-RBD-protein.

Marked virus neutralizing activity was observed as early as week 3 (t2) and rose further by about 6 weeks (t4-t5). However, a decline followed and markedly lower yet positive levels were recorded at around ten weeks after disease onset (t7-t8). Again, patient 2 showed the highest levels.

## Discussion

Our observational study was based on the premise that it would be clinically relevant to investigate the virus load of SARS-CoV-2 along with the development of various antibodies against different regions of the virus, as well as study the virus-neutralizing activity longitudinally. The study was conducted with newly-developed methods in three patients who suffered from a moderately severe course of COVID-19 over a period of about three months.

The clinical course of COVID-19 in these patients turned out to be relatively mild and all three patients recovered after approximately two weeks after symptoms manifested. We were able to detect SARS-CoV-2 RNA over a period of four weeks after onset of symptoms, as evidenced by the significant numbers of RNA genome equivalents in the swabs of the three patients we studied. Even at days 30 and 33 after onset, low amounts of the SARS-CoV-2 genome could be detected in two patients, whereas in the third patient the virus had meanwhile cleared. On days 37, 38 and 40 (Figure 1) complete clearance of the virus was confirmed in all three patients. Our data on the length of time during which SARS-CoV-2 RNA was measurable in the nasal and oro-pharyngeal fluids of patients in this case study inevitably leads to the question about the possible duration of infectivity in post-symptomatic patients. Since the viral load threshold for the transmission of SARS-CoV-2 between humans is yet unknown, it is not possible to predict the impact of close contact between non-immune persons and patients deemed clinically recovered from COVID-19: the policy of social distancing is thus appropriate until complete clearance of the SARS-CoV-2 virus is established. It can well be assumed that SARS-CoV-2 RNA clearance probably indicates the absence of infectivity in recovered SARS-CoV-2 patients.^4,7,8^

In a recent study of hospitalized COVID-19 patients, it was demonstrated that, in nasal and oro-pharyngeal fluid deriving from swabs, high numbers of RNA copies of SARS-CoV-2 could be detected after the onset of symptoms, followed by rapidly decreasing amounts in the subsequent three weeks.^6^ These authors also showed that SARS-CoV-2 from swabs could be isolated in cell culture for up to 8 days after disease onset. By measuring anti-SARS-CoV-2 protein (IgA, IgG) and using a virus neutralization assay, the authors also confirmed that seroconversion had occurred in all patients after 14 days.

It has been postulated that the infection of susceptible human target cells by SARS-CoV-2 occurs via attachment of the receptor binding domain (RBD) of the S1 protein of the virus to the ACE2 receptor of targeted cells, mainly in the respiratory tract.^9,10^ It is assumed that by blocking the attachment of the virus to the receptor, an infection could potentially be inhibited. It was on these grounds that assays were developed for determining human antibodies against this domain of the S1 protein. For diagnostic purposes, however, the detection of antibodies directed against other viral proteins such as the nucleoprotein (N-protein) may also be important, since the N-protein is the most abundant protein in the viral particle.^11^ On the other hand, the immunogenicity of an antibody protein may not only be associated with its nature and amount but also with its effects on the cellular immune response to the virus.^10^

It is common knowledge that, after infections, various immunoglobulins are produced in a certain time sequence. Initially, immunoglobulins of the IgM-class occur and, after several weeks, immunoglobulins of the IgG class appear in blood. While immunoglobulins of the IgA class are also measurable in blood, they also occur in saliva, where their primary role is to counteract infectious agents. We primarily focused on measuring antibodies to SARS CoV-2 Abs of the IgG class. The investigation of IgA antibodies has come into focus more recently, after anti-SARS-CoV-2 IgA antibodies in blood were described to be associated with the early immune response to COVID-19 in patients.^12^ The investigation of IgG and IgA levels against the two SARS-CoV-2 proteins in blood show qualitative similarities but quantitative differences in the patients’ data over time. Interestingly, the patient with the relatively more severe course than the other two patients showed the highest levels of both Abs. This raises two important questions: (1) do antibodies have virus-neutralizing activity and (2) are patients who survive COVID-19 immune to a re-infection in the long term. While it is often the case that anti-viral antibodies are protective, this is not always the case, as is evident after an infection with the Dengue virus. In some instances, the antibodies formed may even have an exacerbating effect on the course of the disease.

We decided to collect information on this subject and also measured the virus neutralizing effect of sera over a period of three months *in vitro*. The course of the *in vitro* virus neutralizing activity in the blood of our patients showed a common pattern: a measurably-high virus load starts soon after the clinical onset of COVID-19, reaching a plateau in the weeks (t3-t6) thereafter, but showing a decline in all individuals after about two months. It is worth noting that serum-neutralizing capacity correlated with the abundance of IgA against the S1-12 RBD in two of the patients. Thus, even though it is clear that the ability of sera to neutralize SARS-CoV-2 depends on several antibody classes, it is possible that S1-specific IgA plays a major role.

Whether our measurements reflect the *in vivo* neutralizing capacity of an individual is yet unclear. However, this finding may indicate that immunity to a re-infection with SARS CoV-2 is probably only transient in nature. Corroborating evidence for this finding must be sought in similar, longitudinal investigations in future, which focus on various indicators of the infectious process during and after recovery from COVID-19 with different clinical outcomes or after vaccination in larger cohorts of patients.

## Data Availability

All data referred in the manuscript are available.

## Acknowledgements

We thank Marija Reichenbächer and Nicole Wizke for their excellent technical assistance. We are also grateful to the patients enrolled in this study for donating blood over the observation period. MS and RB received basic institutional funds from the University Hospital, Tuebingen. Furthermore MS, NR and RB are being supported by grants from the Baden-Württemberg Foundation and the Deutsche Forschungsgemeinschaft (DFG). KK received basic funding by Deutsche Herzstiftung.

